# Self-Reported Mask Wearing Greatly Exceeds Directly Observed Use: Urgent Need for Policy Intervention in Kenya

**DOI:** 10.1101/2021.01.27.21250487

**Authors:** Aleksandra Jakubowski, Dennis Egger, Carolyne Nekesa, Layna Lowe, Michael Walker, Edward Miguel

## Abstract

**Background:** Many countries in sub-Saharan Africa have so far avoided large outbreaks of COVID-19, perhaps due to the strict lockdown measures that were imposed early in the pandemic. Yet the harsh socio-economic consequences of the lockdowns have led many governments to ease the restrictions in favor of less stringent mitigation strategies. In the absence of concrete plans for widespread vaccination, masks remain one of the few tools available to low-income populations to avoid the spread of SARS-CoV-2 for the foreseeable future.

**Methods:** We compare mask use data collected through self-reports from phone surveys and direct observations in public spaces from population-representative samples in Ugunja subcounty, a rural setting in Western Kenya. We examine mask use in different situations and compare mask use by gender, age, location, and the riskiness of the activity

**Findings:** We assess mask use data from 1,960 phone survey respondents and 9,549 direct observations. While only 12% of people admitted in phone interviews to not wearing a mask in public, 90% of people we observed did not have a mask visible (77.7% difference, 95% CI 0.742, 0.802). Self-reported mask use was significantly higher than observed mask use in all scenarios (i.e. in the village, in the market, on public transportation).

**Interpretation:** We find limited compliance with the national government mask mandate in Kenya using directly observed data, but high rates of self-reported mask use. This vast gap suggests that people are aware that mask use is socially desirable, but in practice they do not adopt this behavior.

Focusing public policy efforts on improving adoption of mask use via education and behavioral interventions may be needed to improve compliance.

**Funding:** Weiss Family Foundation, International Growth Centre

## Introduction

When COVID-19 was first detected in the African continent in February 2020, experts feared the pandemic could wreak havoc on low- and middle-income countries (LMICs) with limited health system capacity.^1^ In response, many African countries swiftly adopted strict mitigation strategies that closed borders, restricted movement, and shut markets, schools, and other public places. These measures, along with younger populations and more time spent outdoors, are hypothesized as possible reasons for the relatively low number of recorded COVID-19 cases and fatalities in sub-Saharan Africa.^2,3^ In Kenya, the setting of this study, the official count as of January 11, 2021, stands at 98,271 confirmed cases and 1,710 deaths.^4^ Considerable uncertainty remains about these figures, which are based on targeted testing of symptomatic and high-risk patients.

Masks have emerged as one of the most prominent strategies to reduce SARS-CoV-2 transmission prior to widespread distribution of vaccines, especially in communities where working from home is not feasible and some inter-household interactions are required. Yet compliance with public health guidelines is not guaranteed, and motivating people to adopt new preventive health behaviors is inherently difficult. Quantifying the extent to which mask rules are being followed is essential for setting public health policy decisions. But limited data availability has made understanding the extent to which COVID-19 mitigation rules are being followed challenging. Encouraging mask use in rural, low-income settings with limited ability to absorb economic shocks of lockdowns^5^ and fewer resources to obtain and distribute vaccines^6^ is especially important.

Self-reported mask use estimates in sub-Saharan Africa vary widely,^7-11^ and independent observations of mask wearing is limited to one small scale study that estimated mask use in public transit stations through 45 observations in Ghana.^12^ Further limiting generalizability of the existing evidence is the increased reliance on web-based surveys, which are simultaneously subject to social desirability bias^13^ and selection bias.^14^ These issues are amplified in low-income settings where internet access is not widespread in the general population. Verifying self-reported data with independent observations has been encouraged in health behavior research,^15^ but so far studies measuring mask wearing through wide-scale observations have not been conducted. In Kenya, conflicting anecdotal evidence suggests on the one hand that mask use is strictly enforced^16^ and that compliance is high in informal settlements,^8^ but other reports conclude that access to masks is limited due to insufficient supply.^17^ We use direct observations of public behavior and phone-based surveys to assess mask use in rural Kenya.

## Methods

### Study setting

The first case of COVID-19 in Kenya was detected on March 13, 2020 (Figure 1). Within days, President Uhuru Kenyatta restricted travel from other countries, ordered non-essential government and business employees to work from home, and imposed limits on group gatherings. Schools were closed by March 20^th^ and further restrictions (including a ban of international flights, dusk to dawn curfew, and requirement to end dine-in services) were imposed by March 25th. As in many other countries, the restrictive lockdown measures took a harsh socio-economic toll on the population, including falling incomes, increased food insecurity, and increased domestic violence.^5,18^ In response, lockdowns were gradually eased in favor of promoting the less invasive mitigation strategies of hand washing, mask use, and physical distancing. Starting in early April, masks have been legally mandated in public spaces in Kenya. By July, restrictions on movement in and out of cities were lifted and international flights resumed in August 2020, though curfews remain in effect. The relaxation of travel restrictions has placed greater importance on COVID-prevention measures in rural areas. After multiple waves of COVID-19,^4^ officials in rural settings are becoming increasingly concerned about the spread of the virus there.

**Figure 1.**
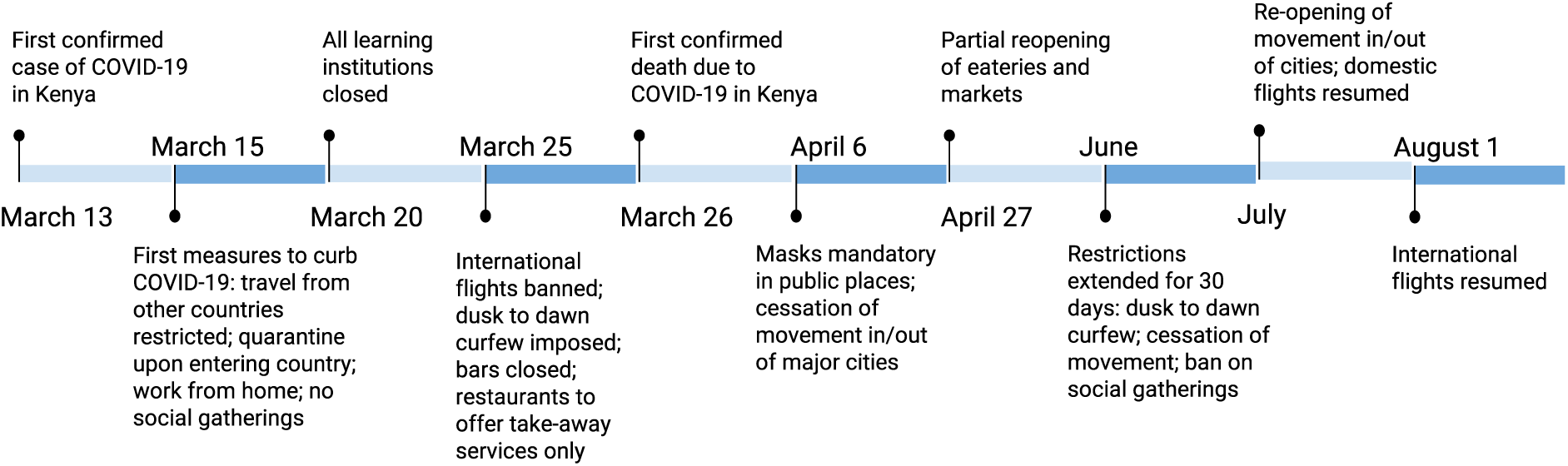
Timeline of COVID-19 in Kenya

### Data Collection Procedures

Data were collected in Ugunja subcounty, located in Siaya County in Western Kenya. This fairly typical rural African setting sits on a major trade route near the border of Kenya and Uganda. We conducted direct observations between August 20 to September 11, 2020, in 71 randomly selected villages and 10 weekly markets. Each village and market were observed for three 1-hour intervals spread over the course of a day. Trained enumerators were instructed to stand at public places and observe behavior related to maintaining physical distancing between people, possession of masks, and proper mask use (i.e., wearing mask over mouth and nose). Common observation places included standing by the side of the road, standing by a shop in the village or between shops in the market, and near public transportation pick-up/drop-off locations. No private information about subjects was collected during direct observations. Public information such as gender, estimated age, activity, and setting type were recorded. The observations were conducted in public spaces without the knowledge of the general public that they were being observed to avoid introducing bias to their behavior.

Household socio-economic surveys were conducted by phone with respondents in Ugunja between June 18 to September 2, 2020. Respondents were randomly selected for participation in the survey from the full set of people living in 166 rural villages in Ugunja, including all 71 villages randomly selected for observations, based on census data collected by members of the research team in late 2019. The surveys gathered information on household characteristics, labor and education, consumption, travel patterns, health, and COVID-19 knowledge and behaviors. We gathered information about masks in the COVID-19 module, including mask ownership, type, and whether respondents wore masks when last visiting public places such their village, markets, religious gatherings, on public transportation, when visiting a shop, or visiting another household. Here, we focus on self-reported mask use in public places that could be compared with direct observations: villages, markets, and public transportation. Expecting some level of bias in self-reports, we also asked respondents to estimate mask wearing behavior of other people by asking them to estimate the proportion of other households in their village wearing masks to the same public places.

The study procedures were approved by the Institutional Review Boards in Kenya (Maseno University) and the United States (University of California, Berkeley).

### Outcome measures

Using direct observations, we constructed three measures of mask use: 1) whether masks were in possession (i.e. visible), 2) whether masks were worn correctly (i.e. covered mouth and nose), and 3) whether no mask was visible to the observers. Using phone surveys, we asked respondents whether they had been to a public place in the past 7 days, and if so, whether they used masks in the specific places they visited. We classify both direct observations and phone survey self-reports by location: within the village, in market centers, on public transport, and other. We then transformed the individual-level phone survey data into person-place observations and constructed three measures of self-reported mask use: 1) always wears masks, 2) sometimes wears masks, and 3) does not wear masks. Respondents who said they never wore masks in public were coded as not using mask in each specific location. Self-reported responses were limited to participants who disclosed they had been to a public place in the week prior to interview. For simplicity, when comparing self-reported and observed data, we group the observed outcomes to a single indicator of whether masks were worn or visible and group the self-reported outcomes to a single indicator of always or sometimes wears masks. We also calculated the estimated proportion of other households wearing masks to the same public places.

### Statistical analysis

We calculated the average mask use in the observed and self-reported samples, stratified by age, gender, and location type. We then used linear regression to test for differences between self-reported vs. directly observed mask use by pooling data across samples and including an indicator for data type. Separately, we also tested whether mask use varied by gender, age, and setting. We explored the direct observations further by grouping observations into three broad categories of activities based on risk of virus transmission: 1) commuting activities, primarily (90%) for-hire motorcycles that require sitting in close proximity to the driver, or bus; 2) solitary activities, including working alone in the field, walking/cycling alone, sitting/resting alone; and social activities, including shopping and talking. Similar measures were not possible to construct with phone data. All linear regression models had standard errors clustered at the village/market and were weighted by the sampling probability of being selected for the phone survey to make reported averages representative of the study area population. We checked the robustness of our findings by restricting the phone survey data to the same dates and the specific villages where observations were conducted.

## Results

We conducted phone interviews with 2,247 respondents from Ugunja subcounty, 87% of whom (1,960 respondents) visited a public place in the 7 days prior to the interview and answered questions about mask use. We observed mask behavior of 9,549 individuals in public places. Approximately 40% of the observed people were women and most were categorized into the two intermediate age categories. Nearly 70% of the phone respondents were women, and again most fell into the two intermediate age categories. Though we randomly selected primary male or female in each household for the survey, more women were available for the interviews. Most observations were conducted in villages. Most phone survey respondents reported visiting markets and walking around their villages.

We document a large discrepancy between self-reported and observed mask use (Table 2). Among the 1,960 respondents who visited a public place in the past 7 days (87% of full survey sample), 76% said they always wore a mask and 12% said they wore a mask some of the time. In direct observations, only 5% of people wore masks correctly and another 5% had masks visible but did not wear them correctly. In other words, while only 12% of people admitted in phone interviews to not wearing a mask in public, 90% of people we observed did not have a mask with them. Self-reported mask use was significantly higher than observed mask use (77.7% difference, 95% confidence interval (CI), 0.742, 0.802). Statistical differences between observed and self-reported mask use persisted when we restricted the samples by gender, age, and location.

**Table 1.** Descriptive characteristics of samples included in direct observations and phone interviews

|  | Observed mask use |  | Self-reported mask use |  |
| --- | --- | --- | --- | --- |
|  | Direct observations (N=9,549) |  | Phone interviews (N=1,960) |  |
|  | N | (%) | N | (%) |
| **Gender** |  |  |  |  |
| Male | 5,482 | (57.4) | 605 | (30.9) |
| Female | 3,825 | (40.1) | 1,355 | (69.1) |
| **Age** |  |  |  |  |
| 19-25 | 2,418 | (25.3) | 40 | (2.0) |
| 26-45 | 5,209 | (54.6) | 767 | (39.1) |
| 46-60 | 1,504 | (15.8) | 384 | (19.6) |
| 60+ | 358 | (3.8) | 303 | (15.5) |
| **Location** |  |  |  |  |
| Market | 1,401 | (14.7) | 1,650 | (84.2) |
| Public transport | 995 | (10.4) | 907 | (46.3) |
| Village | 6,014 | (62.9) | 1,255 | (64.0) |
| Other | 1,139 | (11.9) | 1,100 | (56.1) |

**Notes:** Age and gender in direct observations are estimated by enumerators observing people in public places. Percentages of age and gender may not add to 100% because of missing data: gender was missing for 242 observations and age was missing for 60 observations. Location in phone surveys do not add to 100% because many respondents visited multiple public places in the past 7 days. We categorized the public places into general categories of market centers, villages, public transportation, and “other” places that included religious gatherings, work, and other similar circumstances.

**Table 2.** Statistically higher mask use reported through phone surveys than through direct observations

|  | Observed mask use |  |  | Self-reported mask use |  |  | Self-reported vs. observed mask use |  |
| --- | --- | --- | --- | --- | --- | --- | --- | --- |
|  | Direct observations<br>(N=9,549) |  |  | Phone interviews<br>(N=1,960) |  |  | Always/sometimes<br>vs. worn/visible |  |
|  | Worn | Visible | None | Always | Sometimes | Never | Difference | [95% CI] |
| **Full sample** | 0.047 | 0.057 | 0.896 | 0.759 | 0.117 | 0.124 | 0.772 | [0.742, 0.802] |
| **Gender** |  |  |  |  |  |  |  |  |
| Male | 0.046 | 0.046 | 0.908 | 0.747 | 0.127 | 0.126 | 0.782 | [0.742, 0.821] |
| Female | 0.049 | 0.072 | 0.880 | 0.764 | 0.113 | 0.123 | 0.757 | [0.719, 0.795] |
| **Age** |  |  |  |  |  |  |  |  |
| 19-25 | 0.022 | 0.043 | 0.935 | 0.796 | 0.107 | 0.098 | 0.837 | [0.765, 0.910] |
| 26-45 | 0.051 | 0.057 | 0.892 | 0.765 | 0.118 | 0.117 | 0.776 | [0.737, 0.815] |
| 46-60 | 0.071 | 0.076 | 0.854 | 0.753 | 0.111 | 0.136 | 0.718 | [0.658, 0.777] |
| 60+ | 0.064 | 0.075 | 0.860 | 0.762 | 0.113 | 0.125 | 0.735 | [0.667, 0.803] |
| **Location** |  |  |  |  |  |  |  |  |
| Market | 0.062 | 0.120 | 0.817 | 0.917 | 0.065 | 0.018 | 0.635 | [0.570, 0.699] |
| Public transport | 0.152 | 0.096 | 0.752 | 0.819 | 0.064 | 0.117 | 0.799 | [0.729, 0.870] |
| Village | 0.021 | 0.039 | 0.940 | 0.657 | 0.180 | 0.163 | 0.777 | [0.745, 0.809] |
| **Situation** |  |  |  |  |  |  |  |  |
| Socializing | 0.036 | 0.082 | 0.882 | - | - | - | - | - |
| Alone | 0.032 | 0.049 | 0.919 | - | - | - | - | - |
| Commuting | 0.152 | 0.097 | 0.751 | - | - | - | - | - |

**Notes:** Observed mask use is the proportion of people: 1) wearing masks correctly, 2) having masks visible, and 3) having no mask visible. Self-reported mask use is the proportion of people reporting: 1) always wearing masks, 2) sometimes wearing masks, 3) never wearing masks. We tested for statistical differences in self-reported vs. observed mask use by comparing the proportion of people disclosing they always or sometimes use masks vs. the proportion of people observed wearing or having masks visible using OLS regression with standard errors clustered at the village level, where each row is a separate regression conditional on the descriptive characteristic. Phone data were transformed so that each observation represents a respondent's outing to a public place.

Figure 2 displays the vast discrepancy between self-reported (blue bars) and observed (red bars) mask use by gender, age, and location. Statistical tests results by descriptive characteristics are displayed in Appendix Tables 1 and 2. Self-reported mask use did not differ significantly by gender (Panel A of Figure 2 and Appendix Table 1). Slightly more women had masks visible, but correct mask use was equal among men and women (Panel A of Figure 2 and Appendix Table 2). Mask use increased significantly with the estimated age of observed people (Panel B of Figure 2 and Appendix Table 2), but we found no statistical differences in self-reported mask use by age (Appendix Table 1). Self-reported and observed mask use was significantly lower in villages than in markets or on public transportation (Panel C of Figure 2 and Appendix Tables 1 and 2). Nearly all respondents (98%) reported in phone interviews that they use masks in market centers: 92% said they always wear masks and another 6% said they sometimes wear masks. In comparison, we observed only 18% of people in markets having a mask visibly in their possession, 6% of whom wore masks correctly. A large discrepancy in self-reported vs. observed mask use also existed on public transportation. Notably, the proportion of correct mask use was significantly higher on public transportation (15.2%) than at markets (8.9 percentage point reduction, 95% CI 0.139, 0.040) or in villages (13.0 percentage point reduction, 95% CI 0.167, 0.093).

**Figure 2.**
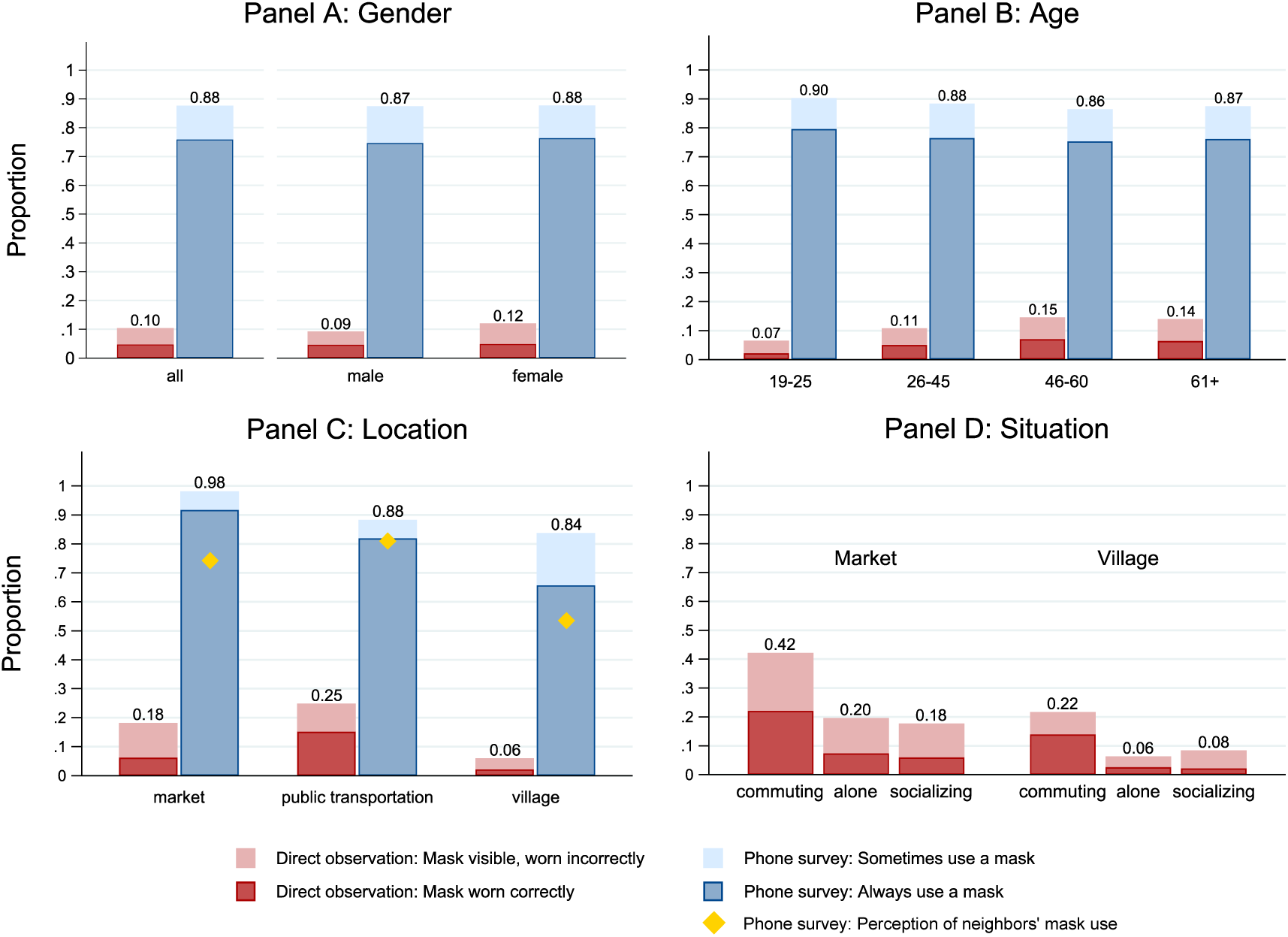
Large discrepancy in self-reported vs. direct observations of mask use in Kenya. Notes: Proportions estimated on sample of 1,960 respondents who had been to a public place in past 7 days and 9,549 observations conducted in 71 villages and 10 market centers. Phone surveys were weighted by the probability of being selected for the phone interviews. Panel B is restricted to participants with age data; no statistically significant differences found in mask use between participants with and without age data. Age and gender in direct observations were estimated by enumerators. Self-reported mask use is significantly higher (p<0.001) than observed mask use in each scenario.

Respondents’ perceptions of mask use by others in their communities were generally lower than their own (diamond markers in Figure 2, Panel C), but were still far higher than direct observations. For example, while 66% of respondents said they always wear masks in villages, their estimate was that about half (54%) of other people in their community wear masks, compared to the 2% of people we actually observed wearing masks correctly. The large discrepancy in mask use via self-reports and estimates of mask use by others compared to the direct observations was evident in all location types.

We also grouped direct observations into three broad activities based on the level of exposure: 1) commuting, 2) solitary activities, and 3) interactive activities. We find that mask use in general was higher in market centers, where there are more interactions with strangers, than in villages (Figure 2, Panel D). Mask use was highest among people who were commuting: in markets 22% wore masks correctly (42% had mask visible) and in villages 14% wore masks correctly (22% had mask visible). We find no meaningful differences in mask use by whether people were interacting or alone.

Since the observations were collected over a shorter time period and a random subset of phone survey villages, we tested robustness of our findings by restricting the phone sample surveys to August to September 2020 in the same 71 villages. We found no meaningful differences in mask use after applying these restrictions (Appendix Tables 3 and 4 and Appendix Figure 1). Age data were missing for 465 phone survey respondents (24% of the sample), but we found no differences in mask use between participants with and without age data (Appendix Table 3).

## Discussion

Masks are among the few tools available to slow the spread of SARS-CoV-2 without imposing major economic hardship, and mask mandates have been implemented or contemplated by governments around the world. Using data from rural Kenyan that is fairly representative of other rural African settings, we find that the strict government COVID-19 mitigation rules regarding mask wearing are adopted to a limited degree. Although only 12% of phone survey respondents admitted to not wearing masks when they visit public places, our observations revealed that the vast majority of people (95%) do not wear masks over their mouth and nose when they are in public. The high proportion of participants who said they wear masks suggests that most are aware that mask use is mandated and socially desirable. However, this awareness has not translated to the behavioral change of adopting mask use in public. This finding suggests that there is an urgent need to improve the adoption of mask use in Kenya and similar settings, via education and behavioral interventions, and possibly increased public distribution of masks.

Although observed mask use was generally low in all of the tested scenarios, we found that people adjusted their mask behavior in more high-risk settings. Mask use was highest on public transportation and in market centers, where interactions with strangers are much more likely. Masks were used correctly (over face and nose) by 22% of commuters in markets and 14% of commuters in villages. In the context of rural Kenya and other similar settings, commuting often involves sharing a motorcycle with a for-hire driver. One possible explanation for the relatively high proportion of commuters wearing masks is that they are more careful to follow public health guidance when they are more at risk. Another explanation is that commuters are especially prone to getting fined for not wearing masks, as random police checks and traffic controls are common in this area.

The existing mask policies in Kenya are enforced with fairly harsh fines ($200 fines have been reported by popular news outlets),^19^ suggesting that further focus on enforcement may have a limited effect. Moreover, enforcement by police may be prone to corruption in this and similar contexts, and in at least one instance a clash over mask use between the police and Kenyan citizens has led to violence and fatalities.^20^ Focusing on other barriers such as lack of access to masks, beliefs about effectiveness of masks or the need to use masks, and behavioral issues related to adopting a new health behavior may be more promising. The vast majority of the phone survey participants in our sample (>99%) reported that they own a mask. However, it is possible that the self-reporting bias we found in mask use was also pervasive in ownership disclosures. Given the relatively low cost of masks in the study setting (approximately$0.50 USD) distribution of free masks may be a worthwhile investment. Pairing mask distribution with public health messages that clearly communicate the risks of COVID-19 and the benefits of masks may be needed to educate the public.^21^ Tailoring the interventions to appeal to social desirability of masks, either through modeling behaviors or stressing altruistic motivations,^22,23^ may be especially relevant in the context of the highly visible behavior of wearing masks, which inherently involves social signaling.

A key methodological takeaway of this study is that measuring mask use through phone surveys may be highly biased. Since the onset of the COVID-19 pandemic, phone and internet surveys have been used increasingly to collect data, both in LMICs and high-income countries.^7,24^ Our findings suggest that studies relying solely on self-reported data to measure adoption of COVID-19 mitigation strategies need to be interpreted with caution. Although our mask wearing estimates using self-reported data are on par with what some other studies report in the literature,^7,8^ direct observations of people in the same communities reveals a vastly different picture of compliance with mask wearing rules. Verifying self-reported data with independent observations may need to be incorporated into public health studies of COVID-19 to provide unbiased estimates of mask use, as has long been encouraged in health behavior research.^15^

Our study was subject to several limitations. Self-reported data, especially when it is related to socially-desirable behaviors, is known to suffer from bias.^13^ Expecting some level of reporting bias, we asked participants not only about their own mask use behavior but also the behaviors of others in the community. Although the proportion of others in the community wearing masks was lower than self-reported mask use, these estimates were still substantial overestimates of the independent observations. Mask use observations were also subject to bias, since we cannot be certain whether people carried masks in a place that was out of sight to the observers.

Nonetheless, since having a mask in a place other than on the face is ineffective in preventing the spread of infection, this bias does not significantly affect our study conclusions. Enumerators were also not able to distinguish whether observed individuals are members of the same household where continuous mask wearing may be impractical. However, since observed mask use was low in *all* settings including public transport and shopping in market centers where people are in close proximity to strangers, this seems unlikely to alter our conclusions. Finally, we acknowledge that the observational nature of the study limits the interpretation of our findings to descriptive statistics that cannot be interpreted as causal.

The progress that the scientific community has made in vaccine development may be slow to translate to widespread distribution in low-income setting such as Kenya, where health systems have limited capacity to supply and distribute even the most basic medicines. With no specific plan for vaccine implementation in sub-Saharan Africa so far developed,^6^ reliance on non-pharmaceutical interventions remains highly relevant for the foreseeable future. Increased focus on encouraging mask use may be needed in cultures where mask wearing is not the social norm.^25^ Mask use remains one of the key public health strategies to reduce COVID-19 transmission in Kenya. Policies to encourage mask use may be especially important in places where shutting down economic activities is harmful to the already low-income population. Our study findings indicate that an increased focus on encouraging mask use via public health campaigns is urgently needed in Kenya and possibly other similar settings in order to close the gap between mask wearing recommendations and actual behavior.

## Data Availability

Data and code available upon request.

## Authors’ contributions

AJ, DE, MW, and EM conceived the study. AJ wrote the first draft of the manuscript. DE, CN, LL, MW, and EM oversaw and implemented data collection. AJ, DE, and LL verified the underlying data, contributed to data analysis, and visualization. All authors contributed to editing of first draft, interpretation of the data, decision to include sensitivity analyses, and editing of the final manuscript. All authors have seen and approve the final text.

