## Appendix for "Self-Reported Mask Wearing Greatly Exceeds Directly Observed Use: Urgent Need for Policy Intervention in Kenya"

Supplementary Appendix

**Appendix Table 1.** Results from OLS regression models testing statistical significance of **self-reported mask use** by gender, age, location

| | Phone surveys | | | | | | | |
|---|---|---|---|---|---|---|---|---|
| | Sometimes uses mask | | | | Always uses mask | | | |
| | coefficient | 95% CI | control mean | N | coefficient | 95% CI | control mean | N |
| **Panel A: Gender** | | | | | | | | |
| *male* | | | 0.874 | 6225 | | | 0.747 | 6225 |
| female | 0.003 | [ 0.036, 0.043] | | | 0.017 | [ 0.031, 0.043] | | |
| **Panel B: Age** | | | | | | | | |
| *19-25* | | | 0.902 | 4759 | | | 0.796 | 4759 |
| 26-45 | -0.019 | [ 0.093, 0.055] | | | -0.031 | [ 0.134, 0.055] | | |
| 46-60 | -0.038 | [ 0.125, 0.048] | | | -0.043 | [ 0.155, 0.048] | | |
| 61+ | -0.028 | [ 0.110, 0.055] | | | -0.034 | [ 0.147, 0.055] | | |
| Joint F-Test | | 0.837 | | | | 0.901 | | |
| **Panel C: Location** | | | | | | | | |
| *public transportation* | | | 0.883 | 3812 | | | 0.819 | 3812 |
| market | 0.099*** | [ 0.074, 0.124] | | | 0.098*** | [ 0.065, 0.124] | | |
| village | -0.045*** | [ 0.069, 0.022] | | | -0.162*** | [ 0.202, 0.022] | | |
| Joint F-Test | | <0.001 | | | | <0.001 | | |

**Notes:** Results from ordinary least squares regression models with standard errors clustered at the village/market level. Each panel represents a separate regression model. Phone data were transformed so that each observation represents a respondent's outing to a public place. Models were estimated separately for each measure of interest: "Sometimes uses mask" and "Always uses mask" using phone data. *P*-value notation: *** p<0.001, ** p<0.01, * p<0.05

**Appendix Table 2.** Results from OLS regression models testing statistical significance of **observed mask use** by gender, age, location, and situation

| | Direct observations | | | | | | | |
|---|---|---|---|---|---|---|---|---|
| | Mask visible | | | | Mask worn correctly | | | |
| | coefficient | 95% CI | control mean | N | coefficient | 95% CI | control mean | N |
| **Panel A: Gender** | | | | | | | | |
| *male* | | | 0.092 | 9300 | | | 0.046 | 9300 |
| female | 0.028** | [ 0.007, 0.049] | | | 0.002 | [ 0.008, 0.013] | | |
| **Panel B: Age** | | | | | | | | |
| *19-25* | | | 0.065 | 9482 | | | 0.022 | 9482 |
| 26-45 | 0.043*** | [ 0.026, 0.059] | | | 0.029*** | [ 0.017, 0.040] | | |
| 46-60 | 0.081*** | [ 0.056, 0.107] | | | 0.049*** | [ 0.033, 0.064] | | |
| 61+ | 0.075** | [ 0.020, 0.129] | | | 0.042* | [ 0.007, 0.077] | | |
| Joint F-Test | | <0.001 | | | | <0.001 | | |
| **Panel C: Location** | | | | | | | | |
| *public transportation* | | | 0.248 | 8394 | | | 0.152 | 8394 |
| market | -0.066 | [ 0.150, 0.018] | | | -0.089** | [ 0.139, 0.040] | | |
| village | -0.188*** | [ 0.248, 0.128] | | | -0.130*** | [ 0.167, 0.093] | | |
| Joint F-Test | | <0.001 | | | | <0.001 | | |
| **Panel D: Situation** | | | | | | | | |
| *socializing* | | | 0.118 | 8404 | | | 0.036 | 8404 |
| alone | -0.037** | [ 0.063, 0.010] | | | -0.003 | [ 0.020, 0.013] | | |
| commuting | 0.131*** | [ 0.073, 0.188] | | | 0.116*** | [ 0.077, 0.156] | | |
| Joint F-Test | | <0.001 | | | | <0.001 | | |

**Notes:** Results from ordinary least squares regression models with standard errors clustered at the village/market level. Each panel represents a separate regression model. Phone data were transformed so that each observation represents a respondent's outing to a public place. Models were estimated separately for each measure of interest: "Sometimes uses mask" and "Always uses mask" using phone data and "Mask visible" and Mask worn correctly" using direct observations. *P*-value notation: *** p<0.001, ** p<0.01, * p<0.05

**Appendix Table 3.** Results from OLS regression models testing statistical significance of self-disclosed mask use by subsets of phone surveys in Ugunja subcounty in Kenya

| | **Phone surveys** | | | | | | | |
|---|---|---|---|---|---|---|---|---|
| | Sometimes uses mask | | | | Always uses mask | | | |
| | coefficient | 95% CI | control mean | N | coefficient | 95% CI | control mean | N |
| *Non-missing age* | | | 0.876 | 6225 | | | 0.761 | 6225 |
| Missing age | 0.004 | [ 0.032, 0.041] | | | -0.016 | [ 0.057, 0.025] | | |
| *Surveyed before August* | | | 0.867 | 6225 | | | 0.738 | 6225 |
| Surveyed August or later | 0.021 | [ 0.016, 0.057] | | | 0.048* | [ 0.006, 0.089] | | |
| *Not in village where direct obs took place* | | | 0.884 | 6225 | | | 0.760 | 6225 |
| In village where direct obs took place | -0.017 | [ 0.055, 0.021] | | | -0.003 | [ 0.045, 0.039] | | |

**Notes:** Results from ordinary least squares regression models with standard errors clustered at the village/market level. Phone data were transformed so that each observation represents a respondent's outing to a public place. Models were estimated separately for each measure of interest: "Sometimes uses mask" and "Always uses mask" using phone data.

**Appendix Table 4.** Statistical differences between self-reports and direct observations persist in models restricted to data from August-September 2020 from the same 71 villages to match timeline and geography of direct observations

| | **Observed mask use** | | | **Self-reported mask use** | | | **Self-reported vs. observed mask use** | |
|---|---|---|---|---|---|---|---|---|
| | Direct observations (N=9,549) | | | Phone interviews (N=) | | | Always/sometimes vs. worn/visible | |
| | Worn | Visible | None | Always | Sometimes | Never | Difference | [95% CI] |
| **Full sample** | 0.047 | 0.057 | 0.896 | 0.772 | 0.089 | 0.139 | 0.757 | [0.699 - 0.814] |
| **Gender** | | | | | | | | |
| Male | 0.046 | 0.046 | 0.908 | 0.789 | 0.075 | 0.136 | 0.772 | [0.664 - 0.879] |
| Female | 0.049 | 0.072 | 0.880 | 0.765 | 0.095 | 0.140 | 0.739 | [0.672 - 0.806] |
| **Age** | | | | | | | | |
| 19-25 | 0.022 | 0.043 | 0.935 | 0.781 | 0.018 | 0.201 | 0.734 | [0.543 - 0.924] |
| 26-45 | 0.051 | 0.057 | 0.892 | 0.818 | 0.115 | 0.068 | 0.825 | [0.782 - 0.867] |
| 46-60 | 0.071 | 0.076 | 0.854 | 0.758 | 0.029 | 0.213 | 0.641 | [0.496 - 0.785] |
| 60+ | 0.064 | 0.075 | 0.860 | 0.717 | 0.113 | 0.170 | 0.690 | [0.542 - 0.839] |
| **Location** | | | | | | | | |
| Market | 0.062 | 0.120 | 0.817 | 0.894 | 0.082 | 0.024 | 0.592 | [0.508 - 0.677] |
| Public transport | 0.152 | 0.096 | 0.752 | 0.806 | 0.035 | 0.159 | 0.794 | [0.718 - 0.869] |
| Village | 0.021 | 0.039 | 0.940 | 0.607 | 0.150 | 0.242 | 0.698 | [0.609 - 0.786] |

**Notes:** Observed mask use is the proportion of people: 1) wearing masks correctly, 2) having masks visible, and 3) having no mask visible. Self-reported mask use is the proportion of people reporting: 1) always wearing masks, 2) sometimes wearing masks, 3) never wearing masks. We tested for statistical differences in self-reported vs. observed mask use by comparing the proportion of people disclosing they always or sometimes use masks vs. the proportion of people observed wearing or having masks visible using OLS regression with standard errors clustered at the village level, where each row is a separate regression conditional on the descriptive characteristic. Phone data were transformed so that each observation represents a respondent's outing to a public place.

**Appendix Figure 1.** Main study findings repeated for a subset of phone surveys conducted during August-September 2020 and the same 71 villages to exactly match the timeline and geography of direct observations data

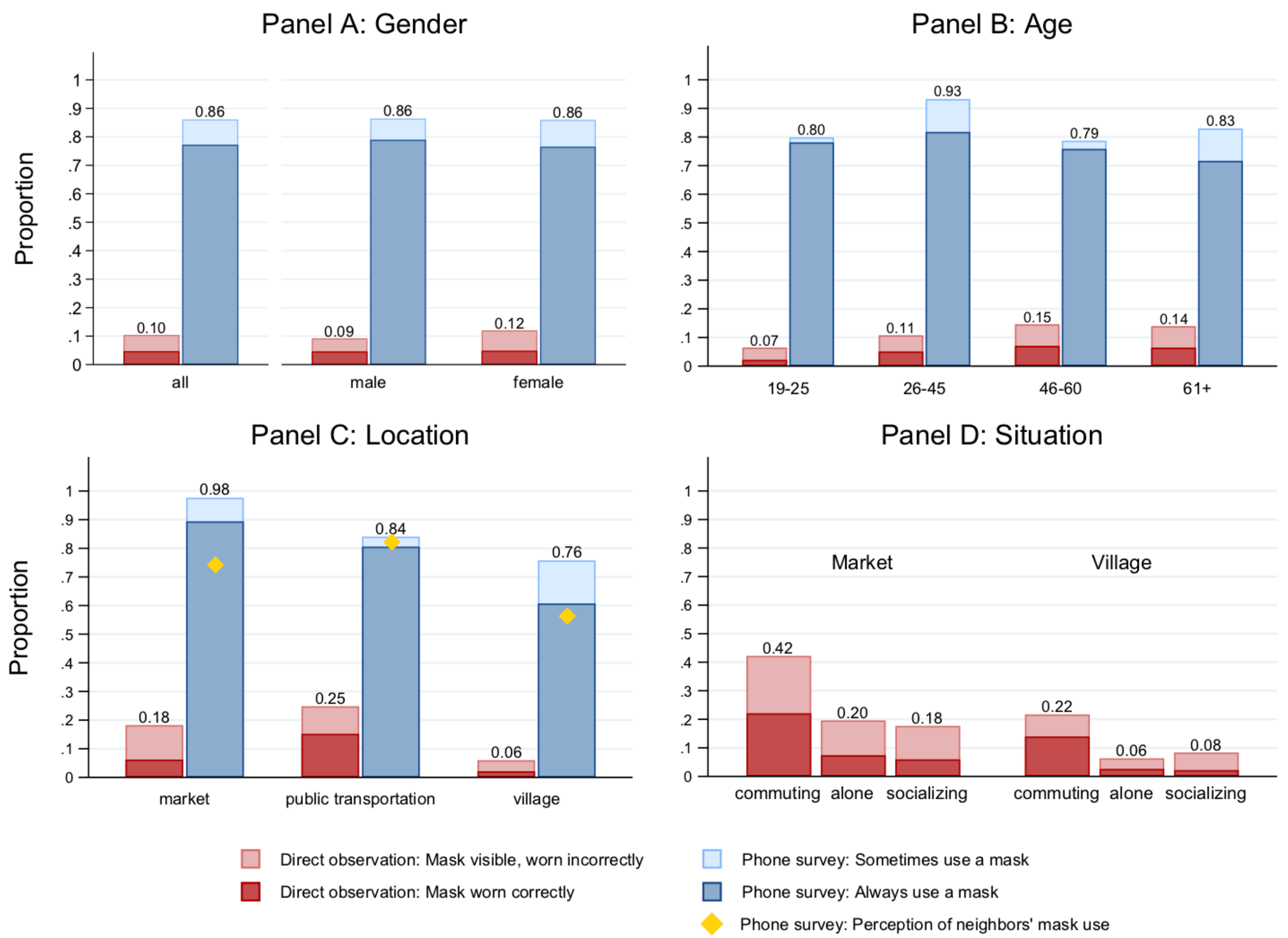


**Notes:** Proportions estimated on sample of 807 respondents who had been to a public place in past 7 days and 9,549 observations conducted in 71 villages and 10 market centers. Phone surveys were restricted to same villages and timeline and weighted by the probability of being selected for the phone interviews. Panel B is restricted to participants with age data; no statistically significant differences found in mask use between participants with and without age data. Age and gender in direct observations were estimated by enumerators.
